# On whether therapeutic plasma exchange is an effective cure against severe/critical COVID-19 pneumonia

**DOI:** 10.1101/2021.04.19.21255657

**Authors:** Luca Cegolon, Behzad Einollahi, Sina Imanizadeh, Mohammad Rezapour, Mohammad Javanbakht, Mohammad Nikpouraghdam, Hassan Abolghasemi, Giuseppe Mastrangelo

**Affiliations:** Local Health Unit N.2 “Marca Trevigiana”, Public Health Department, Treviso, Italy; Nephrology & Urology Research Center, Baqiyatallah University of Medical Sciences, Tehran, Iran; Student Research Committee (SRC), Baqiyatallah University of Medical Sciences, Tehran, Iran; Applied Microbiology Research Center, Systems Biology and Poisonings Institute, Baqiyatallah University of Medical Sciences, Tehran, Iran; Padua University, Department of Cardiac, Thoracic, Vascular Sciences & Public Health, Padua, Italy

**Keywords:** Therapeutic plasma exchange (TPE), COVID-19, clinical and laboratory indexes, cytokine storm

## Abstract

**Background:** There is a risk of novel mutations of SARS-CoV-2 that may render COVID-19 resistant to most of the therapies, including antiviral drugs. The evidence around the application of therapeutic plasma exchange (TPE) for the management of critically ill COVID-19 patients is still provisional and further investigations are needed to confirm its eventual beneficial effects.

**Methods:** We therefore carried out a single-centered retrospective observational non-placebo-controlled trial enrolling 73 inpatients from Baqiyatallah Hospital in Tehran (Iran) with diagnosis of COVID-19 pneumonia confirmed by real-time polymerase chain reaction (RT-PCR) on nasopharyngeal swabs and high-resolution computerized tomography chest scan. These patients were broken down into two groups: Group 1 (30 patients) receiving standard of care (corticosteroids, ceftriaxone, azithromycin, pantoprazole, hydroxychloroquine, lopinavir/ritonavir); and Group 2 (43 patients) receiving the above regimen plus TPE (replacing 2 liter of patients’ plasma by a solution, 50% of normal plasma and 50% of albumin at 5%) administered according to various time schedules. The follow-up time was 30 days and all-cause mortality was the endpoint.

**Results:** Deaths were 6 (14%) in Group 2 and 14 (47%) in Group 1. However, different harmful risk factors prevailed among patients not receiving TPE rather than being equally split between the intervention and control group. We used an algorithm of Structural Equation Modeling (of STATA) to summarize a large pool of potential confounders into a single score (called with the descriptive name “severity”). Disease severity was significantly (Wilkinson rank sum test p-value=0.0000) lower among COVID-19 patients undergoing TPE (median: −2.82; range: −5.18; 7.96) as compared to those non receiving TPE (median: −1.35; range: −3.89; 8.84), confirming that treatment assignment involved a selection bias of patients according to the severity of COVID-19 at hospital admission. The adjustment for confounding was carried out using severity as covariate in Cox regression models. The univariate Hazard Ratio (HR) of 0.68 (95%CI: 0.26; 1.80; p=0.441) for TPE turned to 1.19 (95%CI: 0.43; 3.29; p=0.741) after adjusting for severity.

**Conclusions:** The lower mortality observed among patients receiving TPE was due to a lower severity of COVID-19 rather than TPE effects.

**TRIAL REGISTRATION:** **IRCT registration number:** IRCT20080901001165N58 (Iranian Registry of Clinical Trials)

**Registration date:** 2020-05-27, 1399/03/07 (retrospectively registered)

## INTRODUCTION

COVID-19 is an asymptomatic disease in most cases, but some patients develop life-threatening disease characterized by acute respiratory distress syndrome (ARDS), sepsis, multisystem organ failure (MOF), extrapulmonary manifestations, thromboembolic disease and associated cytokine release syndrome (CRS) [1]. Although the pathophysiology of COVID-19 is far from being completely understood, the severe form of the disease is generally believed to be correlated with over-release of proinflammatory cytokines (tumor necrosis factor, IL-6, and IL-1β), which cause strong inflammation, endothelial injury, thrombotic microangiopathy, multiorgan failure and eventually death [2-4]. The strict biological criteria to diagnose the Cytokine Release Syndrome (CRS) associated with COVID-19 remain however poorly defined [5].

The lack of effective treatments against COVID-19 leads to a sense of urgency to develop new therapeutic strategies based on pathophysiological assumptions, thus endorsing the hypothesis that properly timed anti-inflammatory therapeutic strategies could improve patients’ clinical outcomes and prognosis [6]. The mortality risk associated with the above CRS is thought to increase with the persistence of high blood concentration of cytokines over time, hence some therapeutic strategies against critical COVID-19 are focusing on anti-cytokine treatments or immunomodulators [4,7].

A nonpharmacological option to counteract the dysregulated proinflammatory response featuring severe COVID-19 could be represented by blood purification techniques [6]. Therapeutic plasma exchange (TPE) is an extracorporeal treatment performed by filtrating a volume of plasma equivalent to the estimated plasma volume that selectively removes circulating pathogenetic substances, such as auto-reactive antibodies, immune complexes, paraproteins, lipoproteins, and inflammatory mediators like cytokines. TPE has been applied to manage different critical diseases - including the acute respiratory distress syndrome (ARDS) [8], pneumonia and respiratory failure from H1N1 influenza A virus [9], Kawasaki disease [10] and sepsis - effectively reducing the elevated levels of cytokines and inflammatory mediators, avoiding lethal complications as septic shock, pulmonary embolism, renal injury or disseminated intravascular coagulation [2,8,10-12].TPE as a remedy for the cytokine storm has also already been used as a supportive treatment for critically ill COVID-19 patients [2, 8], especially among those admitted to intensive care unit (ICU) [4].

Overall, TPE, which has been performed for over a century, has proved to be safe and effective in several disorders [13]. However, TPE does not appear in the Coronavirus Disease 2019 (COVID-19) Treatment Guidelines (major revisions on March 5, 2021 and February 23, 2021) issued by the National Institutes of Health (14]. The effect of TPE has not been studied in patients infected with SARS-CoV-1 and MERS.

The risk of novel mutations rendering SARS-CoV-2 resistant to most therapies (including antiviral drugs) might reduce the spectrum of drugs available for COVID-19. TPE can be considered as a salvage or adjunctive treatment against severe COVID-19, with the rationale of clearing out the related cytokine storm and possibly the viral load [15]. The application of TPE in COVID-19 patients is however still not widespread and although limited case reports demonstrated a beneficial effect, but prospective controlled randomized clinical trials (RCT) are necessary to confirm whether this effect could be attributable to TPE administration [16].

In view of all the above, we investigated the effectiveness of TPE in a retrospective controlled non-randomized trial on 73 COVID-19 hospitalized patients at Baqiyatallah Hospital in Tehran (Iran), using mortality as outcome and considering the number and timing of TPE administration along with the severity of illness, ascertained by the heaviest oxygen support required and other relevant risk factors.

## MATERIAL AND METHODS

### Study design

This single-centered retrospective observational controlled - yet not randomized - study enrolled 73 inpatients from Baqiyatallah Hospital in Tehran (Iran),4 March and 20 May 2020. All patients with respiratory symptoms were screened by clinical examination, real-time PCR (rT-PCR) on naso-pharyngeal swabs and chest computerized tomographic (CT) scan to confirm the diagnosis of COVID-19 pneumonia.

### Clinical data collection

The following inclusion criteria were applied to recruit COVID-19 patients for this study:

- age ≥20 years;
- severe pneumonia (defined as tachypnoea (≥30 breaths per min), O2 saturation (SpO2) ≤90% at rest on room air; or PaO2/FiO2 ratio <300 mm Hg);
- positive real-time rT-PCR (Throat-swab specimens) or typical COVID-19 pneumonia imaging at chest CT scan, following WHO interim guidelines: evidence of severe pneumonia, patchy infiltration, ground glass opacities, ill-defined margins, smooth or irregular interlobular septal thickening, air bronchogram, crazy-paving pattern and thickening of the adjacent pleura with severe involvement (> 50%).

The following exclusion criteria were applied [17]:

- non-severe COVID-19 patients,
- MOF in COVID-19 patients.

Patients’ demographic and health data were extracted from medical records. All medical records were screened by double opinion of two hospital doctors. Adjudication of any clinical interpretative diagnostic difference was performed by a pulmonologist. Missing clinical data were filled up by discussion with health care staff.

According to above selection criteria, COVID-19 patients were broken down as shown in figure 1:

**Figure 1.**
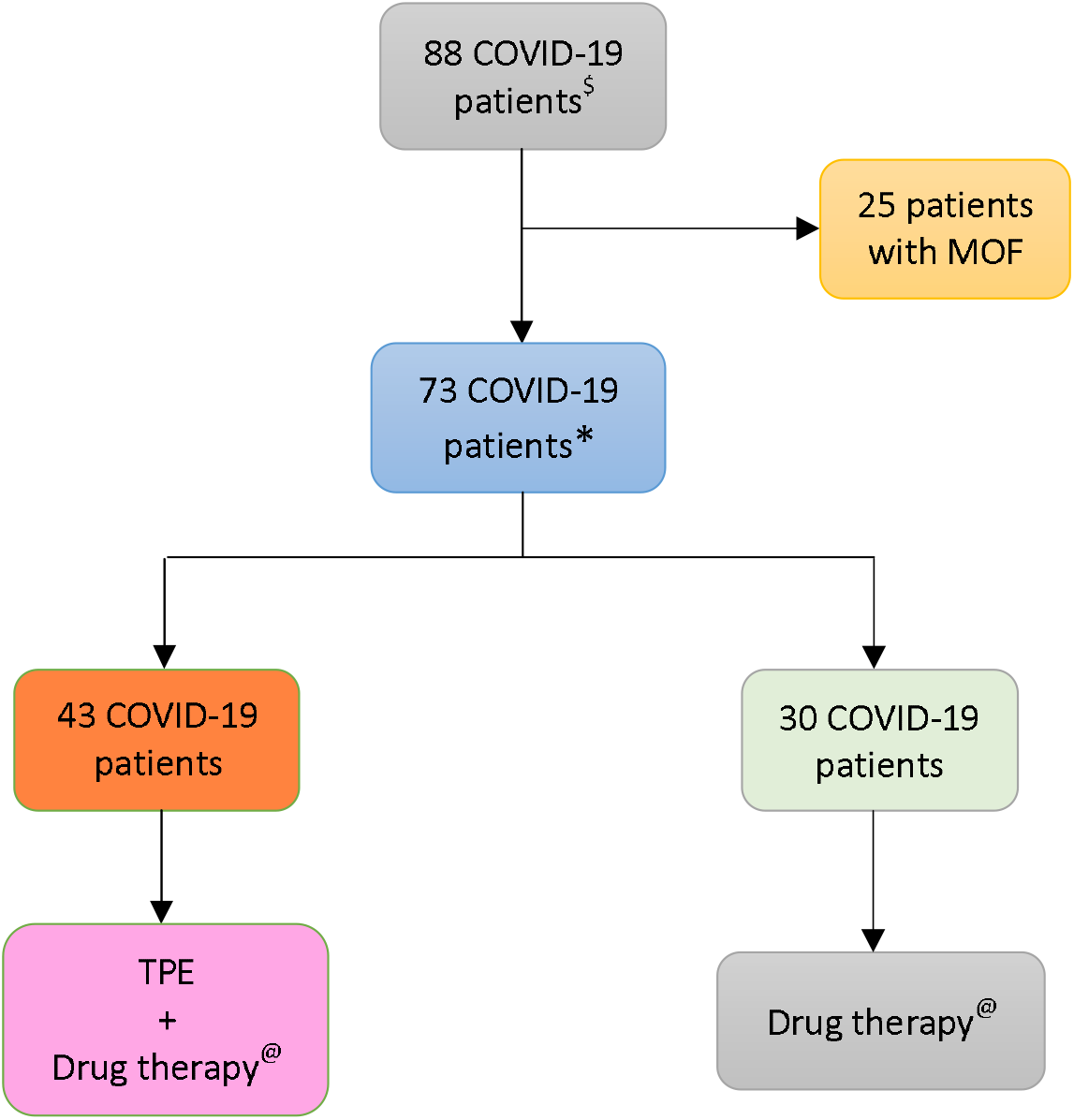
Flow chart displaying the criteria employed to select COVID-19 patients to be treated by therapeutic plasma exchange (TPE). MOF= Multi-organ failure. $ admitted to Baqiyatallah Hospital in Tehran (Iran) between 24 March and 20 May 2020 ∗ - age ≥ 20 years; -severe pneumonia (defined as tachypnoea [≥30 breaths per min], O2 saturation (SpO2) ≤90% at rest on room air; or PaO2/FiO2 ratio <300 mm Hg); -positive real-time RT-PCR (throat-swab specimens); -typical COVID-19 pneumonia imaging at chest CT scan, with patchy infiltration and focal unilateral ground glass opacity, ill-defined margins, smooth or irregular interlobular septal thickening, air bronchogram, crazy-paving pattern, and thickening of the adjacent pleura with severe involvement (> 50%). @ Single daily dose of 500 mg methylprednisolone pulse 250 mg for two days); ceftriaxone (1 gr twice a day); azithromycin (500 mg/daily); pantoprazole (40 mg twice a day); hydroxychloroquine (200 mg/12 hours) and lopinavir/ritonavir (200/50 mg) tablets daily

- **Group 1** (30 patients) received high doses of corticosteroids (methylprednisolone pulse, single daily dose of 500 mg on day 1, 250 mg on day 2 and day3), ceftriaxone (1 gr twice a day), azithromycin (500 mg/daily), pantoprazole (40 mg twice a day), hydroxychloroquine (200 mg/12 hours) and lopinavir/ritonavir (200/50 mg tablets daily).
- **Group 2** (43 patients) receiving the above regimen plus TPE, replacing 2 L of patients’ plasma by 1 L of fresh frozen plasma plasma (FFP) and 1 L of a 5% albumin solution.

### Ethical statement

This study was approved by the Ethical Committee of Baqiyatallah University of Medical Sciences (IR.BMSU.REC.1398435; IRCT registration number: IRCT20080901001165N58; Registration date: 2020-05-27) [17]. All ethical guidelines for studies on human subjects were carefully observed and informed consent was obtained from study participants.

### Variables

#### Risk factors

According to Centers for Disease Control and Prevention [18], older age, chronic obstructive pulmonary disease, cardiovascular disease, type 2 diabetes mellitus; obesity, sickle cell disease, chronic kidney disease, immunocompromised status and cancer are risk factors for severe Covid-19 [18]. We considered these factors if they were actually found in the clinical records of the patients. We also included in the statistical analysis additional conditions for which the data were unclear: co-morbidity (grouping miscellaneous conditions); male sex (that is not currently included on the CDC list of risk factors); type 1 diabetes mellitus; hypertension; and smoking. All the above risk factors were considered dichotomous variables in the statistical analysis.

#### Criteria of classification

In the present study COVID-19 patients were categorized according to death or by TPE administration. For a same risk factor, we estimated the risk of death and/or the probability of being assigned to TPE.

#### Oxygen support

The disease is a severe pneumonia limiting the gas exchange of lung. Rather than chest CT scan imaging, we assumed as indicator of lung involvement the heaviest oxygen delivery support ever administered. The variable was categorized as 0 (high-flow nasal canula), 1 (noninvasive mechanical ventilation) and 2 (invasive mechanical ventilation with intubation). The variable was treated as ordered polytomous variable in the analysis.

#### TPE administration

Early initiation, duration and quantity of TPE could be related to better outcomes. Hence, TPE administration was categorized according to days - ranging from 1 to 12 - of treatment start since hospital admission, coding a new variable (timing) as follows:

- 0 (sample including the above Group 1);
- 1 (patients admitted to TPE on days 1 to 3);
- 2 (patients treated on day 4-5); and
- 3 (patients admitted to TPE 6 to 12 days since hospital admission).

We also coded a variable (n_treat) with 3 levels:

- 0 (including the above Group 1),
- 1 (patients pertaining to Group 2 who underwent 1 to 4 sessions of TPE); and
- 2 (Group 2 patients with 5 TPEs).

#### The latent variable Severity

The latter is not an observed variable but is estimated by SEM program (see below). As can be read in statistical package STATA 14 for SEM analysis, “*a variable is latent if it is not in your dataset but you wish it were. You wish you had a variable recording the propensity to commit violent crime, or socioeconomic status, or happiness, or true ability, or even income. Sometimes, latent variables are imagined variants of real variables, variables that are somehow better, such as being measured without error. At the other end of the spectrum are latent variables that are not even conceptually measurable*”. Severity is a single score summarizing a large number of measured pretreatment covariates that was particularly useful to adjust for confounders using Cox regression models (see below).

### Descriptive analysis

The risk factors of 73 COVID-19 patients, broken down by vital status or TPE treatment, were reported in rows and columns of table 1 to summarize the relationships among observations. At each row and column interception, there were numbers and percentages of subjects having a given trait; the denominator of the percentage was always 53 for patients survived, 20 for those deceased, 43 for patients treated with TPE and 30 for those not undergoing TPE (“Total” in last row of table 1). Risk factors for severe COVID-19 were mainly dichotomic variables (e.g., sex). Table 1 reports only one of the two possible values, the other being easily calculated by subtraction using the total figures (numbers) or 1.00 (percentages). Conversely, all the possible categories of polytomous variables (for example, oxygen support) were reported in table 1. Besides numbers and percentages, table 1 displays the odds ratios (OR), estimated with an exact method due to the relatively limited number of study subjects, with the 95% confidence interval (95%CI) and the two-tail p-value. By default, the conditional maximum likelihood estimates were used in the OR estimation, except for those parameters (e.g., ICU admission) for which a percentage was equal to 100% and the upper bound of 95%CI was infinite. In such a case OR was obtained by Median Unbiased Estimates (MUEs). OR is a measure of association between an exposure and an outcome. The OR represents the odds that an outcome will occur given a particular exposure, compared to the odds of the outcome occurring in the absence of that exposure. The outcome was “death” in the analysis for columns 2 and 3, or “TPE treatment” for columns 5 and 6. The multiple categories of polytomous variables were coded as ordinal variables (0, 1, 2, etc.) but were considered as “*continuous variables*” in the context of exact regression analysis; therefore, only one OR was returned by the statistical program.

**Table 1.**
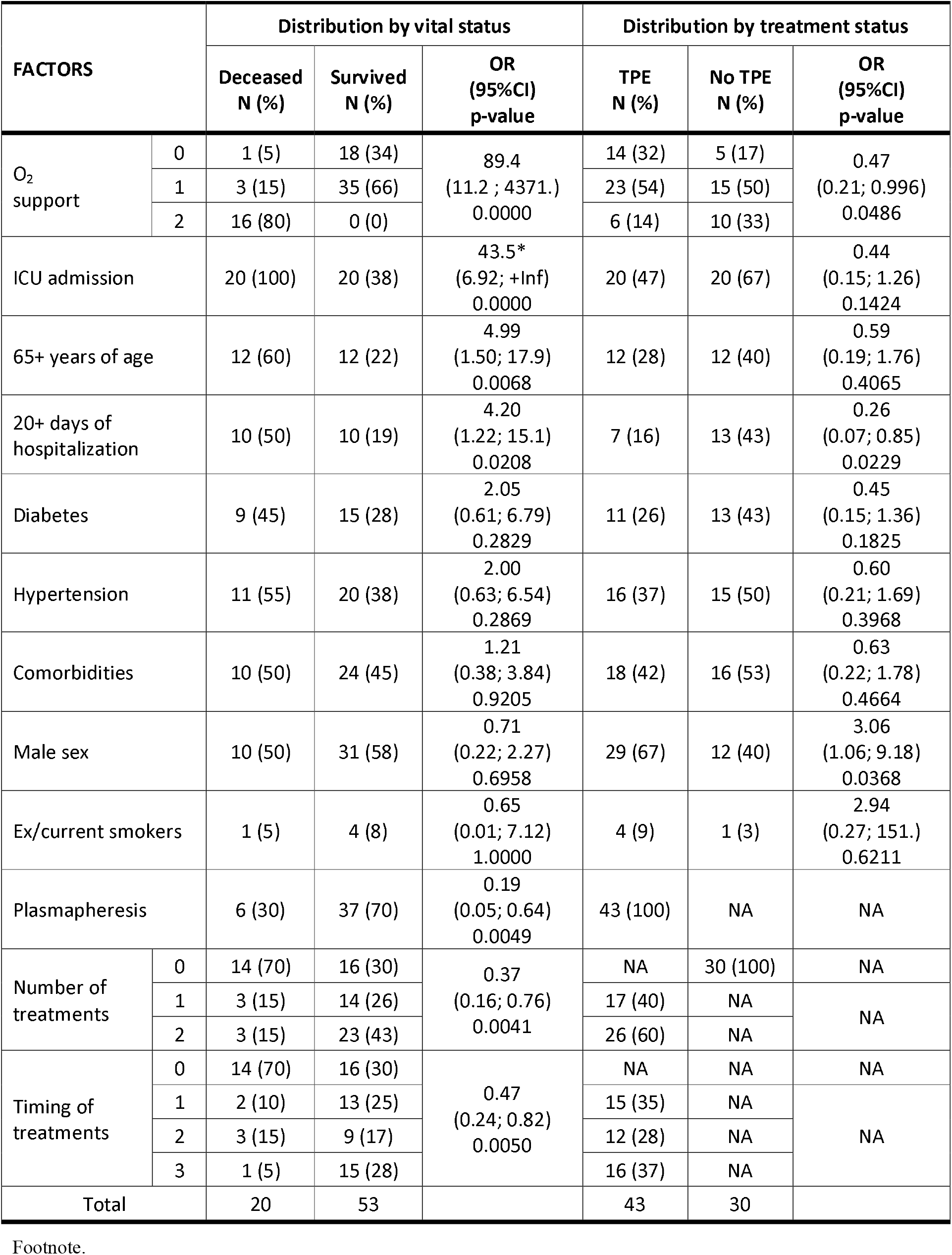
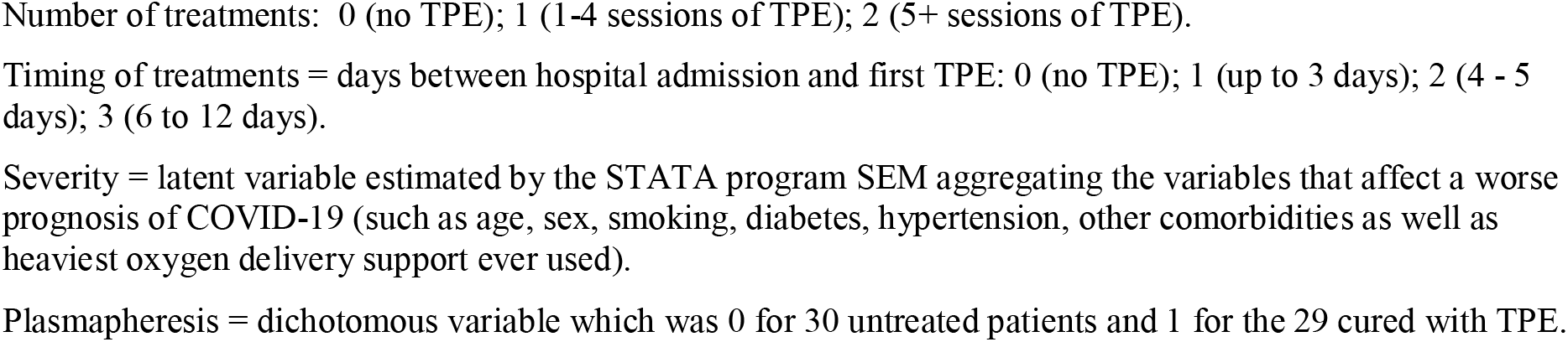
Distribution of 73 COVID-19 patients by vital status and treatment with therapeutical plasma exchange (TPE): number (N) and percentage (%) of cases; exact odds ratio (OR); 95% confidence interval (95%CI) and two-tail p-values.

A numerical value of Severity was estimated by SEM program for each patient. Multiple summary statistics were calculated conditioned on a categorical variable that identified two groups: survived/deceased, or TPE treated/TPE untreated. The numerical statistics for Severity (min, max, median, 25th and 75th percentiles) were reported in table 2, together with the Wilcoxon rank sum test for the equality of the median distribution across the two groups. The distribution of the score Severity score was shown in figure 2 incorporating various vertical lines to mark the median, min and max values of the different samples.

**Table 2.** Distribution of 73 COVID-19 patients by vital status at end of follow-up and by treatment with therapeutical plasma exchange (TPE).

| Latent variable<br>Severity<br>(arbitrary units) | Distribution by vital status |  |  | Distribution by treatment status |  |  |
| --- | --- | --- | --- | --- | --- | --- |
|  | Deceased<br>N=20 | Survived<br>N=53 | Wilcoxon rank-<br>sum test | TPE<br>N=43 | No TPE<br>N=30 | Wilcoxon rank-<br>sum test |
| Lowest value | 5.66 | -5.18 | z = -6.555<br>p = 0.0000 | -5.18 | -3.89 | z = 4.552<br>p = 0.0000 |
| Highest value | 8.84 | -1.27 |  | 7.96 | 8.84 |  |
| 25% percentile | 7.24 | -3.59 |  | -3.61 | -1.73 |  |
| 75% percentile | 8.49 | -2.39 |  | -2.40 | 8.28 |  |
| Median | 7.70 | -2.80 |  | -2.82 | -1.35 |  |

**Figure 2.**
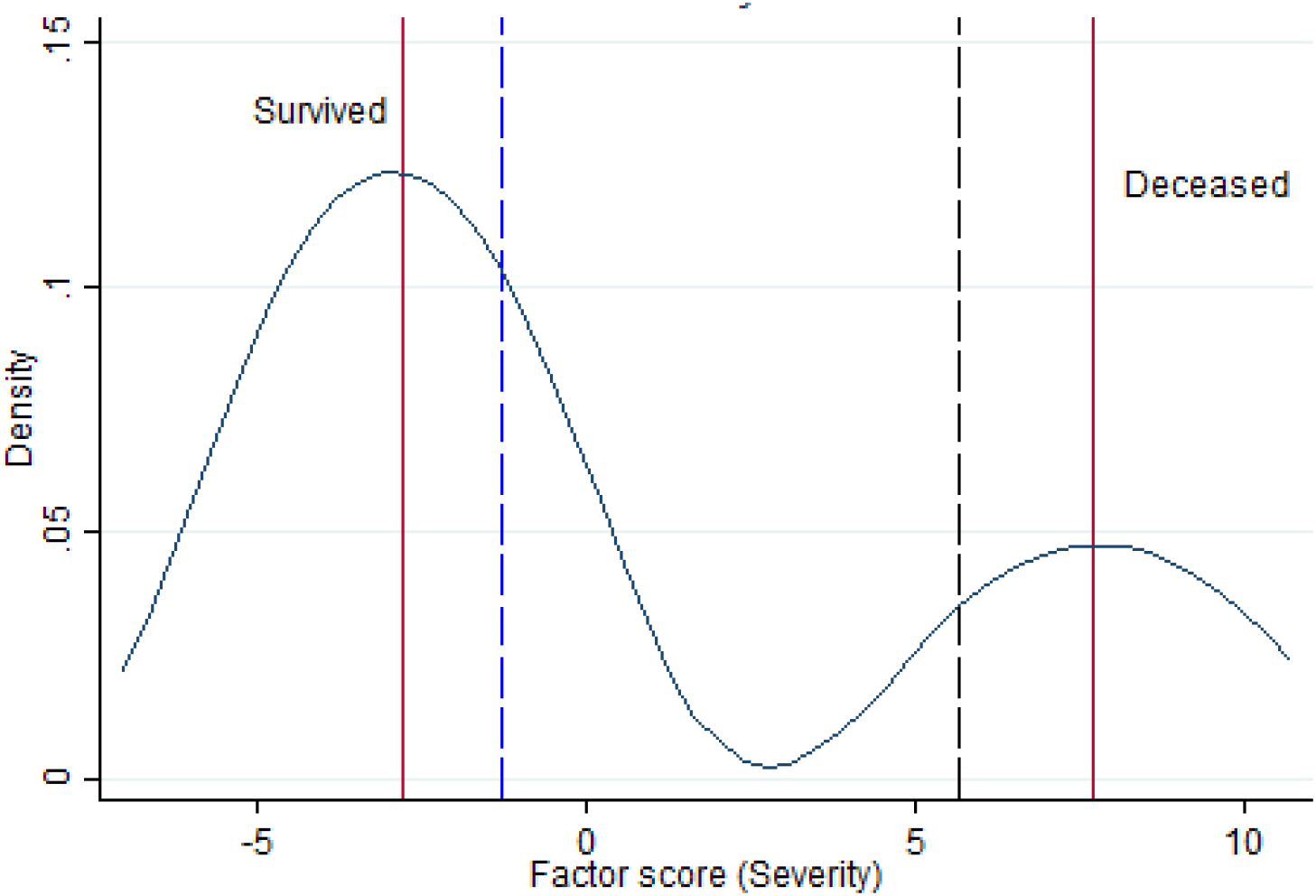
Kernel density estimate for the latent variable “Severity” with vertical lines indicating the median of survived and deceased (red lines), the lowest range of Severity in survivors (blue dashed vertical line) and the highest range of Severity among deceased (black dashed vertical line).

### Statistical analysis for assessing TPE effectiveness

The intervention (TPE) was not randomly allocated to study subjects. To rule out the possibility that any threats were responsible for the observed treatment effect, we used a conceptual framework based on knowledge of the relevant literature. We contrasted the two central aspects of the study: TPE therapy (including number and timing of administration) and the latent variable Severity, using mortality as outcome.

All the above assumption were converted into a SEM model. The STATA command syntax for the model was:

*sem (Severity -> age sex smoking diabetes hypertension co_morbidities oxygen_support) (mortality <-Severity tpe n_treat timing), stand vce(oim)*

“Severity” is capitalized because SEM program commands assume that variables are latent if the first letter of the name is capitalized. In the first and second set of parentheses we specified, respectively, the estimations of the latent variable “Severity” (i.e., identification of a plausible confounder) and the model for the final outcome (i.e., adjustment by illness severity of the mortality associated with number and timing of TPE treatment, and treatment as a whole). In STATA commands, “stand” specifies that the effects are expressed as standardized (or beta) coefficients that make comparisons easy by ignoring the independent variable’s scale of units, while “vce(oim)” is the default and specifies how the standard errors are calculated. We used two SEM goodness-of-fit statistics: (1) the chi square test for “model versus saturated” (the saturated model is the model that fits the covariances perfectly); and (2) the coefficient of determination (CD) that is like R^2^ for the whole model, a perfect fit corresponding to a CD of 1. SEM results were both tabulated and presented graphically (see figure 3).

**Figure 3.**
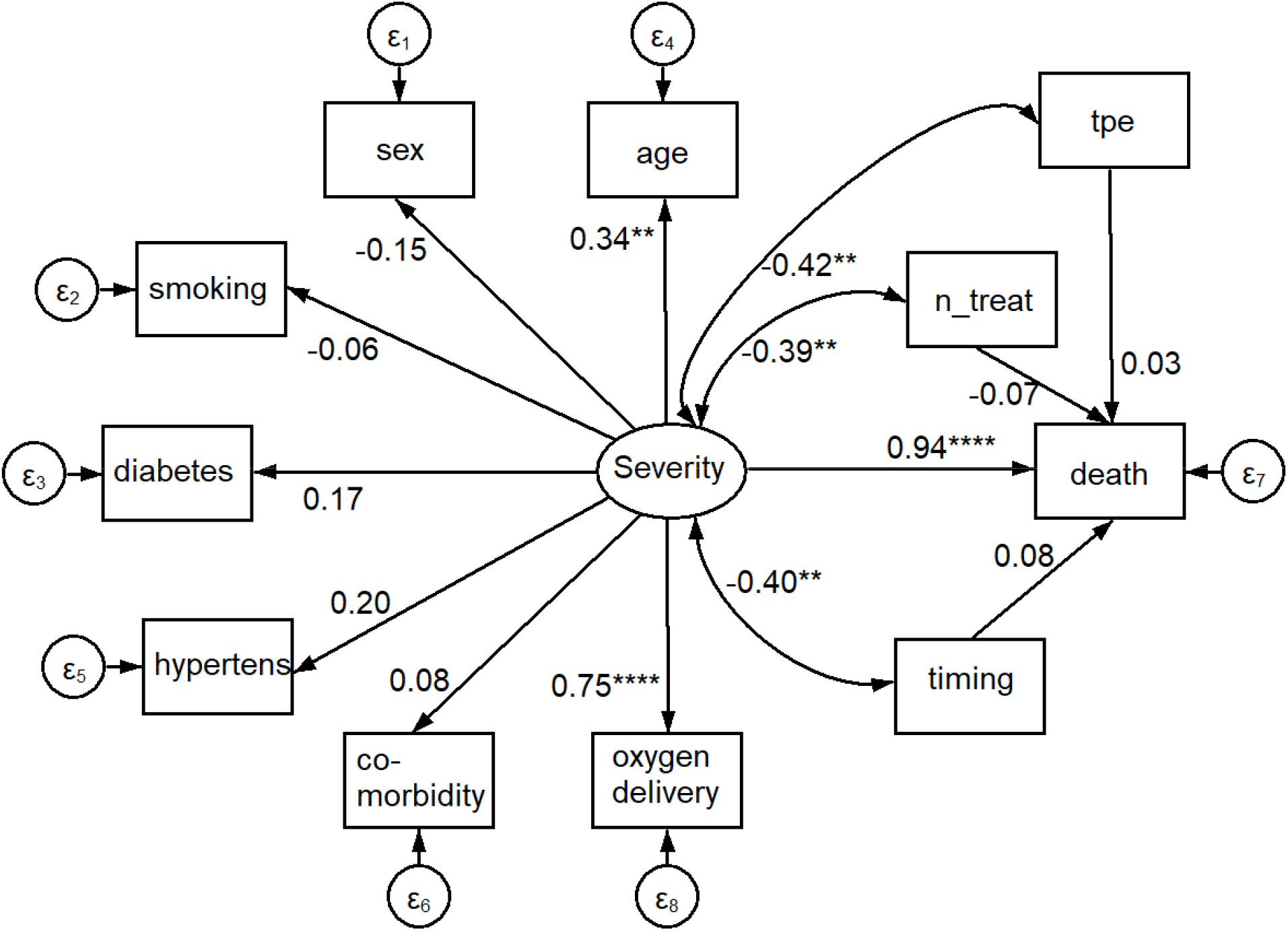
Path diagram of results shown in Table 3. An oval indicates the latent variable (Severity, square boxes indicate the observed variables, circles indicate errors, arrows specify the direction of causal flow, an arrowed route is a path, curved paths express covariances, and the estimated beta coefficients appeared along the paths. It can be seen that: Severity is significantly associated with O2 delivery (p=0.000) and age (p=0.002); there is a negative correlation between Severity and plasmapheresis (“tpe”, p=0.003), number (“n_treat”, p=0.006) and timing (“timing”, p=0.005) of its administration; the impact on mortality (the dependent variable “death”) was highest for Severity (p=0.000) and close to 0.0 for plasmapheresis variables.

The sample size required for SEM is dependent on model complexity. The best option is to consider the model complexity (i.e., the number of exogenous variables) and the following rules of thumb: minimum ratio 5:1, with a recommended ratio of 10:1, or a recommended ratio of 15:1 for data with no normal distribution [19]. With four exogenous variables (tpe, n_treat, timing, Severity) used in the SEM model, we should have a minimum of 20 (= 4 × 5) to a maximum of 60 (= 4 × 15) subjects; in total we reached 73 subjects with complete data, thus fulfilling these requirements.

**Table 3.**
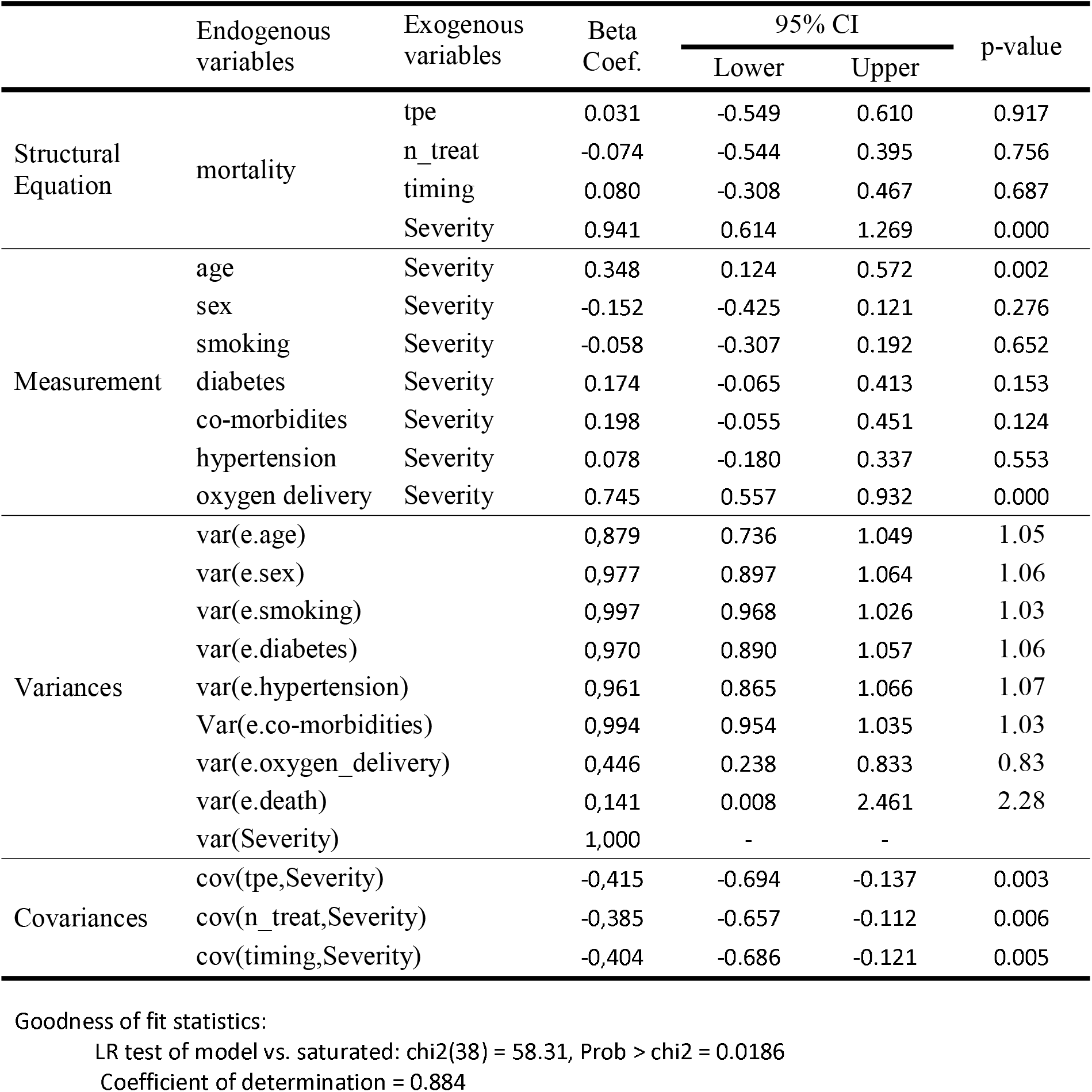
Four groups of SEM results (structural equations, measurement, variances, covariances) for the analysis of mortality of critically ill COVID-19 patients after therapeutic plasma exchange. Standardized beta coefficients (with “minus” sign indicating inverse relationship) with lower and upper limit of 95% confidence intervals (95%CI) and p-values; SEM’s goodness-of-fit statistics at bottom of table.

Furthermore, to examine how the above factors influenced the rate of mortality happening at a particular point in time, the survival analysis using the Cox proportional-hazards models was adopted. Since the test “rho” of proportional-hazards assumption was not statistically significant for each covariate and the global test was neither statistically significant (data not shown), we therefore started by computing univariate Cox analyses for all variables (overall TPE therapy, n_treat, timing, severity), then we fitted multivariate Cox analysis using various models (with different numbers of covariates) to describe how the factors jointly impacted on survival.

Assuming a HR of 0.4, the sample size for Cox proportional-hazard model was estimated to be 38.

Complete case analysis was adopted including all 73 patients. In all analyses 0.05 was set as threshold of statistical significance. All analysis were conducted with the statistical package STATA 14.

## RESULTS

### Descriptive results

Table 1 reports under the heading “Distribution by vital status” the factors of interest in decreasing order of OR value. O2 support was the most important factor associated with death from COVID-19; worth of notice that no patient survived to invasive mechanical ventilation with intubation. Admission to ICU and hospitalization longer than 20 days came as second and fourth relevant factors, but they were believed collinear variables associated with oxygen support and no longer considered. Older age significantly increased five times the risk of death. Diabetes and hypertension were found to double the risk of death but the impact was not statistically significant probably because of the few cases involved. Co-morbidity (other than the above-mentioned disease) appeared to increase only slightly the risk of death. Male gender and smoking did not play any role on mortality. The dichotomous variable “plasmapheresis” and the polytomous variables “number of TPE” and “timing of TPE” consistently showed an important and highly significant protective effect, suggesting TPE as effective intervention protecting from death among critically ill COVID-19 cases.

Data of the same patients, in the same order of risk factors, were reported in the last three columns of table 1, under the heading “Distribution by treatment status”. Here the point of view was whether the different harmful exposures were split equally between the intervention and control group or prevailed somewhere. Table 1 shows that whenever an OR in the fourth column was higher than unity it was lower than unity in the seventh column, suggesting that damaging factors prevailed among untreated patients.

Deaths were 6 (14%) in Group 2 and 14 (47%) in Group 1. However, the protecting effect of TPE could be due to some confounding characteristics of groups being compared rather than TPE treatment itself. Low frequencies and sparse data did not allow detecting a meaningful key of interpretation. Therefore, the problem was analyzed with SEM, estimating the latent variable “Severity”.

The score test Severity, based on arbitrary units, had negative values indicating lower severity and positive values suggesting higher severity of illness. As can be seen in figure 2 Severity had a bimodal distribution with local maximal values of two modes corresponding to the median value of survived and deceased patients (red vertical lines in left graph). The graph of figure 2 also shows that the lowest range of Severity in survivors (−1.18, blue dashed vertical line) did not overlap with the highest range of Severity among deceased (5.66, black dashed vertical line).

Table 2 shows that the median Severity was 7.70 in survivors and −2.80 in non-survivors, a highly significant difference (z = −6.555; p = 0.0000). Moreover, the rank sum test of Wilcoxon proves that the Severity medians (equal to −2.82 in treated patients and −1.35 in untreated patients) in treated and untreated groups were significantly different (z=4.552; p=0.000), even though the distributions were overlapping, confirming that selection for treatment involved a selection of patients for the initial severity of COVID-19.

### Outcome results

SEM results are:

- Structural equations. This includes the beta coefficients (with a “minus” sign indicating an inverse relationship), 95% confidence intervals (95%CI) and p-values for each of two structural equation models.
- Measurement. The standardized (beta) coefficients for this measurement model can be interpreted as correlation coefficients describing the direction (positive or negative) and degree (strength) of relationship between each indicator and the latent variable PAH.
- Variances. The variability explained (1 – error) by the above fitting
- Covariances. SEM uses a set of concurrent regression equations to yield coefficient estimators and the covariance is a measure of cross-equation correlation.

Table 3 shows that, according to results of “Structural Equations”, death was due to Severity of disease (beta coefficient = 0.91; 95%CI: 0.63 to 1.20; and p=0.0005), not to number or timing of TPE treatment. As shown in “Measurement” the latent variable Severity was mainly correlated with oxygen delivery methods (beta coefficient = 0.75; 95%CI: 0.57 to 0.93; p=0.0005) and age (beta coefficient = 0.40; 95%CI: 0.16 −0.44; p=0.005), while a non-significant borderline result (p=0.061) was observed for diabetes. The findings in the section “Covariances” demonstrated that Severity of disease was negatively correlated with TPE as a whole (beta coefficient = −0.42; 95%CI: −0.69; −0.134; p=0.003); number of treatment (beta coefficient = −0.39; 95%CI: −0.66 to −0.11; p=0.006) and timing of TPE (beta coefficient = −0.40; 95%CI: −0.69 to −0.12; p=0.005), showing that treated subjects had initially a lower severity of disease. The seemingly protective effect of TPE shown in Table 1 was therefore a selection bias.

Using the graphical interface of SEM, the same results of Table 3 were displayed as path diagram in figure 3. In this figure, square boxes stand for variables, arrows specify the direction of causal flow, an arrowed route is a path and the cross-equation correlation is displayed as a curved path. The estimated beta coefficients with corresponding p-values appeared along the paths. The figure is a useful synthesis of the findings. It can be seen that: Severity is significantly associated with O2 delivery (p=0.000) and age (p=0.002); there is a negative correlation between Severity and plasmapheresis (“tpe”, p=0.003), number (“n_treat”, p=0.006) and timing (“timing”, p=0.005) of its administration; the impact on mortality (the dependent variable “death”) was highest for Severity (beta coefficient = 0.94; 95%CI: 0.61; 1.27; p=0.000) and close to zero for plasmapheresis variables.

Table 4 shows the results of Cox proportional-hazard models, where quantities called hazard ratios (HR) measure the impact (i.e., the effect size) of covariates. When a HR is significantly higher than unity (p = 0.007), severity decreases the length of survival and is a bad prognostic factor. The univariate HR of 0.68 (95%CI: 0.26; 1.80) for TPE turned to 1.19 (95%CI: 0.43; 3.29) after adjusting for severity. The increasing HR for TPE administration going from univariate to multivariate analyses can be attributed to a confounding effect of disease severity; i.e., the lower mortality observed among patients receiving TPE was due to a lower severity of their disease rather than TPE effects. The same interpretation applies to the parallel changes of number of TPE and timing of its administration.

**Table 4.**
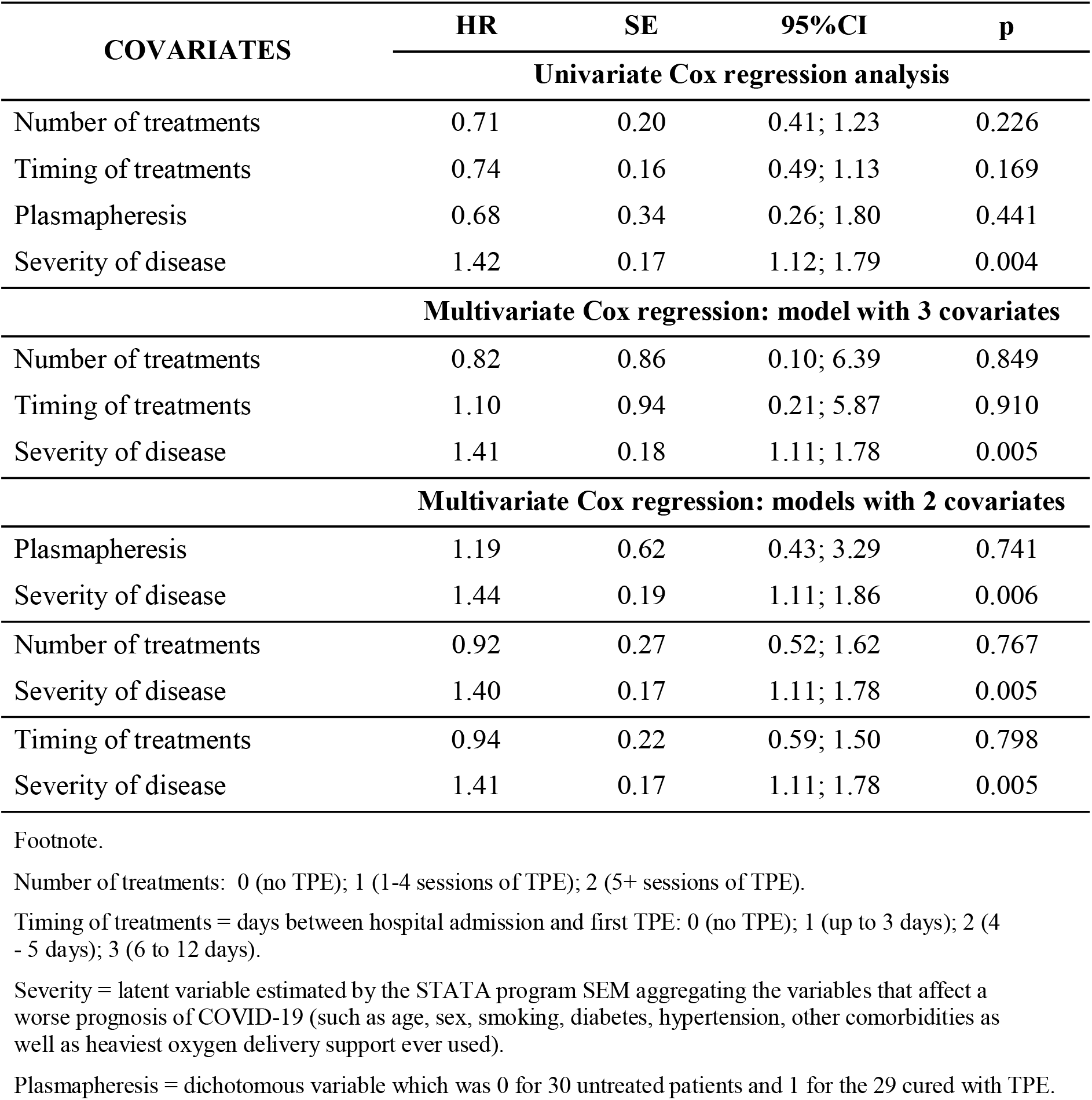
Effect of therapeutic plasma exchange (TPE) on 30-day mortality of 59 patients hospitalized for severe COVID-19 pneumonia. Results of univariate and multivariate Cox proportional-hazards models: hazard ratio (HR), standard error (SE), 95% Confidence Interval (95%CI) and p-value (p). One model with 3 and three models with 2 covariates (description in footnote).

## DISCUSSION

### Key findings

This single-centered retrospective observational non-placebo-controlled trial enrolled 73 inpatients from Baqiyatallah Hospital in Tehran (Iran) with diagnosis of COVID-19 pneumonia confirmed by real-time polymerase chain reaction on nasopharyngeal swabs and high-resolution computerized tomography chest scan. These patients were broken down into two groups: Group 1 (30 patients) receiving standard care (corticosteroids, ceftriaxone, azithromycin, pantoprazole, hydroxychloroquine, lopinavir/ritonavir); and Group 2 (43 patients) receiving the above regimen plus TPE (replacing 2 liter of patients’ plasma by a solution, 50% of normal plasma and 50% of albumin at 5%) administered according to various schedules. The time window of observation was 30 days and all cause mortality was the outcome. Deaths were 6 (14%) in Group 2 and 14 (47%) in Group 1. We used an algorithm of Structural Equation Modeling (STATA software) to summarize a large pool of covariates into a single score (called with the descriptive name of “severity”). Disease severity was significantly (Wilkinson rank sum test p-value=0.0000) lower among TPE treated (median: −2.82; range: −5.18; 7.96) compared to patients not treated with TPE (median: −1.35; range: −3.89; 8.84). The adjustment for confounding was carried out using severity as covariate in Cox regression models. The univariate HR of 0.68 (95%CI: 0.26; 1.80; p=0.441) for TPE turned to 1.19 (95%CI: 0.43; 3.29; p=0.741) after adjusting for severity. Thus, the lower mortality observed among patients receiving TPE was due to a lower severity of their disease rather than TPE effects.

### Limitations

All patients underwent measure of creatinine, blood urea nitrogen, white blood cells, lymphocytes, neutrophils, hemoglobin, and platelet the day of hospital admission and on day 3, day 6, and last day of hospitalization; whereas levels of c-reactive protein (CRP), lactate dehydrogenase, creatine phosphokinase, erythrocyte sedimentation rate, alanine aminotransferase and aspartate transaminase were assessed only the first and last day of hospital stay. By contrast, ferritin, d-dimers, serum il-6 level, SOFA score and APACHE II score were not available. The latter parameters, together with PiO2/FiO2, oxygen saturation, respiratory rate, lymphocyte count, neutrophil–lymphocyte ratio, CRP and lactate dehydrogenase, have critical cutoffs for decision making onto whether initiating TPE in COVID-19 patients according to a group of physicians from different parts of the world with extensive expertise in clinical apheresis and critical care [20]. Therefore, the diagnosis of CRS could be uncertain. However, the lack of data cannot be a major limitation because, on one hand, assessing all parameters may not be necessary to start, stop or refrain from TPE and single or multiple combinations of these parameters can be used to assess eligibility for TPE and subsequent responses [20]; and, on the other hand, all 73 patients were affected by severe COVID-19 pneumonia with evidence of hypoxemia (respiratory rate > 30/minute or partial pressure of oxygen on arterial blood gas less than 80 mmHg or PaO2/FiO2 less than 300) and lung infiltrates on more than 50% of the lung field, according to WHO criteria [21].

### Interpretation

Illness Severity as a single summary score of a large number of covariates, a key topic of the present study, was included in the statistical analysis to achieve several advantages.

- First, confounding adjustment using regression methods, particularly nonlinear models such as Cox regression models, requires sufficient number of outcome events (roughly 10 outcome events per covariate) [22]. For example, to adjust for 5 confounders, one would need to have 5×10 = 50 outcome events. In our study, however, outcome events, i.e., deaths, were only 20 (table 1).
- Use of a summary score rather than a large number of measured pretreatment covariates avoids over-fitting and collinearity issues in estimating treatment effects [23].
- An alternative method to estimate a single summary of all covariates is propensity score matching (PSM). If we find two individuals with the same propensity score, one in the treated group and one in the untreated group, we can assume that these two individuals are more or less “randomly assigned”, i.e., equally likely to be treated by TPE or not with respect to measured pretreatment characteristics [24]. However, during matching procedure, the closest untreated and treated individuals are matched and the remaining untreated individuals are excluded from the analysis [24]. Our study avoids exclusion of unmatched individuals because the summary score Severity was included as a covariate in a regression model on treatment, where mortality (outcome variable) was regressed against TPE (treatment variable) and Severity (representing any threats for the observed effect) (table 3 and 4).

As shown in table 3, Severity was correlated with maximum O2 delivery support (beta coefficient=0.75; 95%CI=0.56-0.93); p=0.000); the higher the respiratory support, the higher the severity of disease, which in turn was the only determinant of 30-day mortality in our patients affected by severe COVID-19 pneumonia. Deng et al [25], analyzing the clinical characteristics of survivors versus non-survivors of COVID-19 in Wuhan (China), found that O2 saturation level among non-survivors (O2 sat: 85%; range: 77%-91%) compared to survivors (O2 sat: 97%; range: 95%-98%) was significantly lower (Z = 10.625; p < 0.001). Another striking finding was that the lowest range of O2 saturation range in survivors (95%) did not overlap with the highest range of O2 saturation among non-survivors (91%) [25]. The latter findings agree with those of figure 2, showing that the lowest range of Severity in survivors (−1.18, blue dashed vertical line) did not overlap with the highest range of Severity among deceased (5.66, black dashed vertical line). On the other hand, in a randomized controlled clinical trial on the use of TPE in patients with life-threatening COVID-19 [5] HR for the partial arterial pressure of oxygen to fractional inspired concentration of oxygen (PaO2/FiO2 ratio) was 0.98 (95%CI = 0.96-1.00) with p-value = 0.02 at Cox proportional hazards multivariable regression model using 35-day mortality as outcome.

It has been reported that in patients affected by severe COVID-19 the cytokine storm was significantly higher around 7–14 days after the disease onset [26]. Thus, timely initiation of plasmapheresis within this period could determine better health outcomes. It is also necessary to administer TP for the correct duration and quantity to monitor the possible drug removal of specific therapies and to follow the treatment outcome [27]. The effect of plasmapheresis in sepsis has shown that both the timing and disease severity are important for the beneficial effect of the procedure [28]. Therefore, we coded two variables, one for number of treatments administered (n_treat) and another for timing of the first administration (timing). Along with TPE treatment as a whole, these two variables were included in SEM analysis, all of them showing to be negatively correlated with Severity (see “Covariances” in table 3). All of them had no effect on survival of COVID-19 patients since the corresponding HR was not significantly different from 1.00 (table 4).

### Generalizability

The current evidence on the effect of TPE in severe/critical COVID-19 was summarized by Lu [29]. Among the 24 studies reviewed, 21 had a non-experimental design (case-reports or case series without controls), in which the effectiveness of the intervention had been assessed by before-and-after comparison of findings. TPE protocols were quite heterogenous, the treatment schedules ranging from one to nine procedures, usually administered daily, but sometimes every other day. Fresh frozen plasma was specified as the replacement fluid for many, but not all studies. The duration of the procedure and plasma volume exchanged were also variable. On the basis of these studies cannot be concluded that TPE is a potential treatment for SARS-CoV-2 infection. According to Honore et al. [30], “*we are unable to clearly identify at the bedside which patients are in a pro-inflammatory state that could kill them or an anti-inflammatory state that could help them to survive” and therefore “it is currently impossible when employing an unselective removal technique to know if we are doing something good by removing an excess of pro-inflammatory mediators or something wrong by removing anti-inflammatory mediators … that could help patients to survive … TPE also has the potential to cause harm by diluting or attenuating the protective antibodies formed by the patient to infection and depletion of immunoglobulins and complement components 3 and 4*”. The same topics along with new comments are reported in another letter of Honore et al [31]: “*Reduction in the plasma level of inflammatory mediators via the use of TPE does not necessarily equate to an improvement in the septic status of the patient. It is simply an artificial reduction, treating the numbers, so to speak. … the improvement can be rather due to the various additional treatments that the patients received* “. Lastly, according to Stahl et al [32] “*we would be very cautious in recommending TPE … as the procedure itself might remove critically important neutralizing antibodies against SARS-CoV-2. We gained further insight that we would like to share, when we recently performed rescue TPE in a life-threatening situation of a septic COVID-19 patient. We could not only detect SARS-CoV-2-specific IgG and IgA antibodies in the waste bag plasma but did also reduce the circulating amount of antibodies by one log step*.*”*.

Out of 24 studies reviewed by Lu [29], 3 had a quasi-experimental design, using two groups of COVID-19 patients, either treated or untreated with TPE, in order to disentangle the effects of TPE from those of additional treatments. A no significant difference in all-cause mortality was reported by two [33, 34] of three studies. The third study [35] is discussed below together with a similar study [36] found by us.

In both the latter studies, treatment assignment was a non-random procedure and pretreatment characteristics of groups were not comparable. Nonetheless, direct comparison between treated and untreated groups was carried out, finding a significant difference in mortality (p=0.037) in the study of Gucyetmez [35] and in time for CRS resolution (p=0.04) in the study of Kamran [36]. These findings could be explained by either the treatment, or pretreatment variables, or both. In order to form matched groups of treated and untreated individuals with similar or comparable pretreatment characteristics, the propensity score matching (PSM) was applied to both Gucyetmez [35] and Kamran [36] studies. After PSM, the two matched groups (TPE and non-TPE) each of 45 patients [36] or 12 patients [35] had comparable pretreatment characteristics and hence differences in the respective outcome became more significant (p=0.001 in the former and p=0.009 in the latter study). However, while in randomized controlled trial (RCT), randomization ensures comparability in both measured and *unmeasured* pretreatment characteristics, in PSM the comparability of the treatment groups is limited only to measured pretreatment characteristics included in the propensity score model. In the study of Gucyetmez [35] there were 18 patients treated and 35 patients untreated, becoming 12 in each group after PSM. In Kamran [36] study the corresponding figures were 71 (TPE), 209 (no-TPE) and 45 (after PSM). During matching, the closest untreated and treated individuals are matched and the remaining individuals that were not matched are excluded from the analysis. Exclusion of unmatched individuals from the analysis not only affects the precision of the treatment effect estimate but also could have consequences for the generalizability of the findings [37].

The last study [5], not examined by Lu [29] but found by us, is a prospective randomized controlled trial (experimental design) based on the comparison of 43 TPE treated patients with 44 patients receiving only the standard of care, all randomly selected among COVID-19 patients admitted to ICU. The use of randomization gives the experimental design greater strength. We can be more certain that any differences between the intervention group and the control group, with respect to the apparent effect of the intervention, can be attributed to the intervention, and not to group differences. The statistical analysis using Cox regression models demonstrated that TPE did not significantly affect 35-day mortality, agreeing with the results of our present study.

## CONCLUSIONS

In absence of sufficient data on proven efficacy, the usefulness of TPE procedure in COVID-19 appears questionable in the light of the considerations drawn by Patidar [20]:

- *“TPE is an expensive, time- and resource-intensive procedure not readily available everywhere, especially in rural settings and low- and middle-income countries”*.
- *“Adverse events of TPE (e*.*g*., *citrate toxicity, hypotension) may contribute to hemodynamic instability”*.
- *“Removing patient’s neutralizing antibodies or therapeutic drugs during TPE can possibly delay recovery or reduce therapeutic benefit”*.
- *“Using albumin or saline as a replacement fluid can increase risk of coagulopathy”*.
- *“Instruments require decontamination if used for COVID patients or requirement of dedicated apheresis instrument for treatment of COVID-19 patients”*.

## Data Availability

The datasets generated and analysed during the current study are not publicly available, since they were purposively collected by the authors for the present study, but are available from the corresponding author on reasonable request.

## Ethics approval and consent to participate

This Ethical Committee of Baqiyatallah University of Medical Sciences approved this study (IR.BMSU.REC.1398435; IRCT registration number: IRCT20080901001165N58; Registration date: 2020-05-27). All ethical guidelines for studies on human subjects were carefully observed and informed written (??) consent was obtained from study participants.

## Consent for publication

Not applicable.

## Data Availability Statement

The datasets generated and analyzed during the current study are not publicly available, since they were purposively collected by the authors for the present study, but are available from the corresponding author on reasonable request.

## Authors’ contributions

LC and GM designed and run the analysis, interpret the data and drafted the original draft; BE, MJ, SI, MR, MN, HA conceived the study, collected the data, contributed to interpret the data and validated the manuscript.

## Acknowledgements

None.

